# Sampling of the Lung Microbiome in Patients Undergoing Lung Resection

**DOI:** 10.64898/2026.08.22.26360295

**Authors:** Alexander Pohlman, Andrew D Marten, Melline Fontes Noronha, Mark Khemmani, Alan J Wolfe, Zaid M. Abdelsattar

## Abstract

**Background:** Although the lung is of low biomass, it harbors a diverse and dynamic microbiome that may influence disease and healing. Existing studies have used diverse sampling methods with high propensities for contamination and sampling error, leading to diverse and unclear results. Here, we characterized the lung microbiome via airway and parenchymal samples to determine variation across patients and sampling methods.

**Methods:** We recruited adult patients undergoing lung resection for suspected or confirmed malignancy. After resection and under sterile conditions, a 1 cm cubic piece of non-cancerous lung parenchyma and a swab from the specimen’s bronchus were collected and sent for microbiome analysis via 16S rRNA gene amplicon (V4) sequencing on an Illumina platform. An established bioinformatics pipeline was used to determine taxonomic identification. Baseline clinical and demographic data were compared to microbiome composition.

**Results:** A total of 86 patients were included in the study. Beta diversity (microbial composition) varied significantly by sampling method (biopsy of lung parenchyma versus airway swabs), so all further results were analyzed within sample types. Further analyses revealed significant differences in beta diversity by lobe of the lung, indicating a different microbial composition by anatomic location. Analyses of patient demographics revealed significant differences by age and comorbidities, including chronic obstructive pulmonary disease and atrial fibrillation.

**Conclusions:** The lung harbors a diverse microbiome that differs by anatomic location and patient characteristics. This study provides a framework for more accurate future lung microbiome sampling and characterization.

## Introduction

Lung resections are commonly performed for biopsy and removal of suspected or confirmed lung cancer.^1^ Increasing evidence suggests the microbiome can play a role in cancer development and surgical outcomes.^2–4^ Studies have shown significant variation in lung microbiome composition both within and across studies, with some showing associations with various respiratory conditions and patient-specific factors.^5^ Existing literature suggests that thoracic surgical operations also can alter the lung microbiome. For example, physiologic shifts associated with one-lung ventilation are reported to shift microbial abundance.^6^

Despite increasing recognition of microbial influences on cancer and surgical outcomes, ongoing studies to establish lung microbiome roles are limited by small sample sizes, diverse and unreliable sampling techniques (*e.g.*, bronchoalveolar lavage [BAL]), or lack of clinical context.^7–11^ The current sparse literature reports various microbes within the lung with poorly understood effects, in part due to patient variation and a knowledge gap concerning optimal sampling strategies.

Here, we characterized the lung microbiome of patients undergoing lung resection to determine microbial diversity across patients and identify associations with clinical and demographic characteristics. We hypothesized that within each participant, the parenchyma, sampled by biopsy, and the airways, sampled by a swab, would harbor different microbiomes. We further hypothesized that, within the parenchyma, the microbiome would differ by anatomic location. This study provides evidence for future sampling methodologies for improved study of the lung microbiome and its association with cancer pathogenesis and outcomes.

## Materials and Methods

### Patient Population

In this prospective study, we recruited and consented adult patients with suspected or confirmed lung malignancy undergoing surgical resection at our institution. Resection types included lobectomy, segmentectomy, and wedge resections. Our institutional review board approved this study (IRB#215744). All indications for surgery and type of lung resection were determined by the operating surgeon per standard practice. All patients received standard surgical prophylactic antibiotics within one hour of incision.

### Sampling Technique

Both parenchyma (biopsy) and airway (swab) samples were obtained for microbiome analysis. After resection, under sterile conditions, a 1cm cubic piece of non-cancerous lung parenchyma from the edge of the resected sample was sharply cut and placed in a sterile container for the parenchymal sample. For anatomical resections, the bronchus was sharply cut, and a white-capped Eswab^®^ (Becton Dickinson, Franklin Lakes, NJ) was used to sample the inside of the bronchus and placed into a sterile collection solution. Samples were placed in a biohazard bag, kept at 4°C, and brought directly to the research laboratory, where a nucleic acid preservative (10% AssayAssure, Sierra Molecular, Incline Village, NV) was added to all samples and stored at −80°C. Samples then underwent DNA extraction and sequencing **(Supplemental Figure 1)**. To assess potential DNA contamination, extraction negative controls (no parenchyma or swab) were processed with the samples.

### Bioinformatic and Statistical Analysis

From the raw sequence reads, adapters were trimmed. Low-quality reads were removed using Cutadapt.^12^ DADA2 software processed the remaining high-quality reads (*i.e.,* filtering, dereplication, chimera removal, construction of an amplicon sequence variant (ASV) abundance table). To achieve taxon identification, BLCA^13^ was used to query NCBI and Greengenes2 databases.^14^ For each ASV, identification with confidence at the lowest taxonomic level was used for downstream analyses. ASVs with taxa confidence scores less than 69.5% were assigned “unclassified/unknown.” Data is accessible at NCBI BioProject: ID#PRJNA1428797, entitled “Pulmonary Oncogenesis Microbiome Following Surgery.”

*Lysobacter* ASVs were removed as they are known contaminants of the enzyme achromopeptidase used for genome extraction. Low abundance ASVs were removed. Contaminant ASVs were determined via a scoring method that encompasses results from Decontam,^15^ nonparametric statistical testing (Kruskal-Wallis p≥0.5), mean comparison (*i.e.*, mean counts of samples versus extraction controls), and whether sample read counts were less than 5x greater than controls.

Alpha (within-sample) and Beta (between-sample) diversity indices were computed using statistical packages phyloseq^16^ and vegan^17^ in R (R Foundation for Statistical Computing, Vienna, Austria). Definitions of diversity measures can be seen in **Supplemental Table 1**.

Medians with interquartile range (IQR) were presented for alpha diversity measures and within-sample comparisons of differences were performed using Wilcoxon signed rank tests for comparisons between 2 groups or Kruskal-Wallis for comparisons between 3 or more groups. Beta diversity results were compared by abundance-based Bray-Curtis and presence/absence-based Jaccard indices. The analysis included correction for multiple comparisons and statistical significance was determined by Permutational Multivariate Analysis of Variance (PERMANOVA). DESeq2 was used to identify taxa enriched in one variable relative to another ^18^

## Results

Ninety patients were consented for inclusion. Eighty-six patients had microbiome specimen analyses completed. Of these, the average age was 68.1±9.3, 50 (58.1%) were female, 77 (89.5%) were white, and 65 (75.6%) were smokers. There were 58 (67.4%) lobectomies, 24 (27.9%) wedge resections, and 4 (4.7%) segmentectomies. Operations were most commonly completed minimally invasively by either robotic-assisted surgery (84.9%) or video-assisted thoracoscopic surgery (VATS;12.8%). The most common histologies were adenocarcinoma (58.1%) and squamous cell carcinoma (19.8%; **Table 1)**. Three participants had only biopsy samples and no swabs due to logistic reasons.

**Table 1.** Study population demographics Demographic^1^ Study Population Age,mean±SD 68.1±9.3.

| **Demographic<sup>1</sup>** | **Study Population** |
| --- | --- |
| **Age**, mean±SD | 68.1±9.3 |
| **Sex**, Female | 50(58.1%) |
| **Race** |  |
| White | 77(89.5%) |
| Black | 6(7.0%) |
| Asian | 3(3.5%) |
| **Ethnicity**, Hispanic | 12(14.0%) |
| **BMI**, mean±SD | 29.3±6.3 |
| **Medical History** |  |
| Diabetes | 23(26.7%) |
| Chronic Obstructive Pulmonary Disease | 29(33.7%) |
| Coronary Artery Disease | 14(16.3%) |
| Chronic Kidney Disease | 7(8.1%) |
| Congestive Heart Failure | 6(7.0%) |
| Hypertension | 54(62.8%) |
| Cerebrovascular Accident | 7(8.1%) |
| Atrial Fibrillation | 11(12.8%) |
| Prior Cardiothoracic Surgery | 9(10.5%) |
| **Pulmonary Function Tests**, mean±SD | |
| FEV1 | 83.1±16.8 |
| DLCO | 75.8±18.6 |
| **Pre-operative Steroid Use** | 11(12.8%) |
| **Smoker** |  |
| Current | 14(16.3%) |
| Former | 51(59.3%) |
| Never | 21(24.4%) |
| **Smoking History**, pack-years | 27.3±25.0 |
| **Lobe** |  |
| Right Upper | 27(31.4%) |
| Right Middle | 8(9.3%) |
| Right Lower | 17(19.8%) |
| Left Upper | 17(19.8%) |
| Left Lower | 17(19.8%) |
| **Histology** |  |
| Adenocarcinoma | 50(58.1%) |
| Squamous | 17(19.8%) |
| Other | 19(22.1%) |
| **Operation** |  |
| Lobectomy | 58(67.4%) |
| Segmentectomy | 4(4.7%) |
| Wedge | 24(27.9%) |
| **Surgical Approach** |  |
| Robotic | 73(84.9%) |
| VATS | 11(12.8%) |
| Open | 2(2.3%) |

<sup>1</sup>SD—standard deviation, BMI—body mass index, FEV1—forced expiratory volume in one second, DLCO—diffusing capacity of the lungs for carbon monoxide, VATS—video-assisted thoracoscopic surgery

To compare microbiomes of lung parenchyma (biopsy) and large airways (swab), we constructed a histogram presenting the top 15 genera by relative abundance (**Figure 1A**). Cross-sectionally, the two samples did not differ by alpha diversity (**Figure 1B)**; thus, the two sample types did not differ by richness (the number of taxa), evenness (the distribution of taxa), or relative abundance. However, the two samples differed significantly by beta diversity (Bray-Curtis Dissimilarity Index PERMANOVA p=0.008; Jaccard Index PERMANOVA p<0.001; **Figure 1C**). DESeq2 analysis revealed three ASVs that differed significantly between the sample types: ASV9 (*Corynebacterium*) and ASV52 (*Pseudomonas*) were enriched in biopsy samples; ASV50 (GWA2-37-10) was enriched in swab samples (p<0.001; **Figure 1D**). We conclude that biopsy and swab samples differ significantly. Thus, for subsequent analyses, we analyzed each sample type separately.

**Figure 1.**
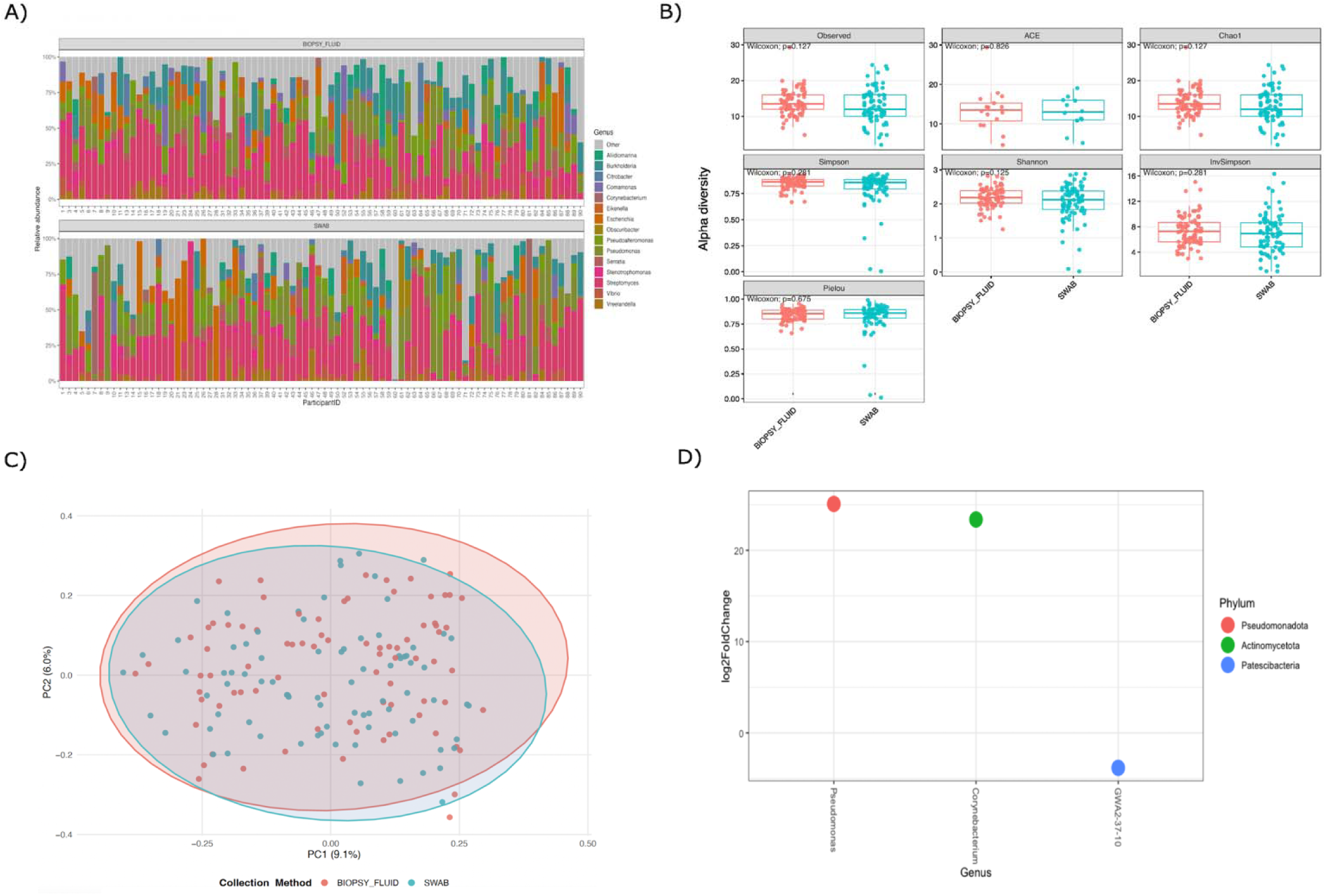
Comparison of microbial diversity by sampling method: biopsy versus swab. (A) Relative abundance of the top 15 genera found in biopsy and swab samples; all other genera are included as “other.” (B) Comparisons of alpha diversity by richness (Observed, Abundance-based coverage estimator [ACE] and Chao1), by evenness (Pielou) and by composites of richness, evenness, and relative abundance (Simpson, Shannon, Inverse Simpson). (C) Principal Coordinates Analysis (PCoA) based on the Bray-Curtis dissimilarity index, revealing significant differences in beta diversity (PERMANOVA p=0.008). (D) DESeq2 Analysis revealing enriched species in different sample types. Positive Log_2_FoldChange enriched in biopsies; negative Log_2_FoldChange enriched in swabs.

The microbiomes differed significantly by participant (biopsy: Bray-Curtis PERMANOVA p<0.001, Jaccard PERMANOVA p=0.004; swab: Bray-Curtis PERMANOVA p=0.016, Jaccard PERMANOVA p=0.004; **Supplemental Figure 2**). In contrast, parenchyma and airway microbiomes did not differ by several variables, including surgeon, sex, race, ethnicity, BMI, smoking history, and steroid use (all PERMANOVA p>0.05).

We also sorted participants dichotomously by age (40-59 versus 60-89 years); older participants exhibited a more diverse biopsy microbiome by overall alpha diversity measures (Simpson, Shannon, and Inverse Simpson; Wilcoxon p=0.036, 0.035, and 0.036, respectively), primarily due to increased richness (Observed and Chao1; Wilcoxon p=0.054; **Figure 2A**). Beta diversity analysis indicated a significant difference in composition (Bray-Curtis PERMANOVA p=0.053; **Figure 2B**). DESeq2 analysis revealed 7 ASVs that differed significantly between age groups. ASV21 (an unknown member of the phylum Pseudomonadota), ASV23 (*Corynebacterium*), ASV35 (*Citrobacter*), ASV40 (*Burkholderia*), ASV44 (unknown member of Pseudomonadota), and ASV46 (*Stenotrophomonas*) were enriched in older participants; ASV360 (*Aeromonas*) was enriched in younger participants (all p<0.001; **Figure 2C**). In contrast, alpha and beta diversity analyses indicated no significant difference by age in swabs (PERMANOVA p>0.05; **Supplemental Figure 3A-B**). We conclude the parenchymal microbiome differs by age, but airway microbiome does not.

**Figure 2.**
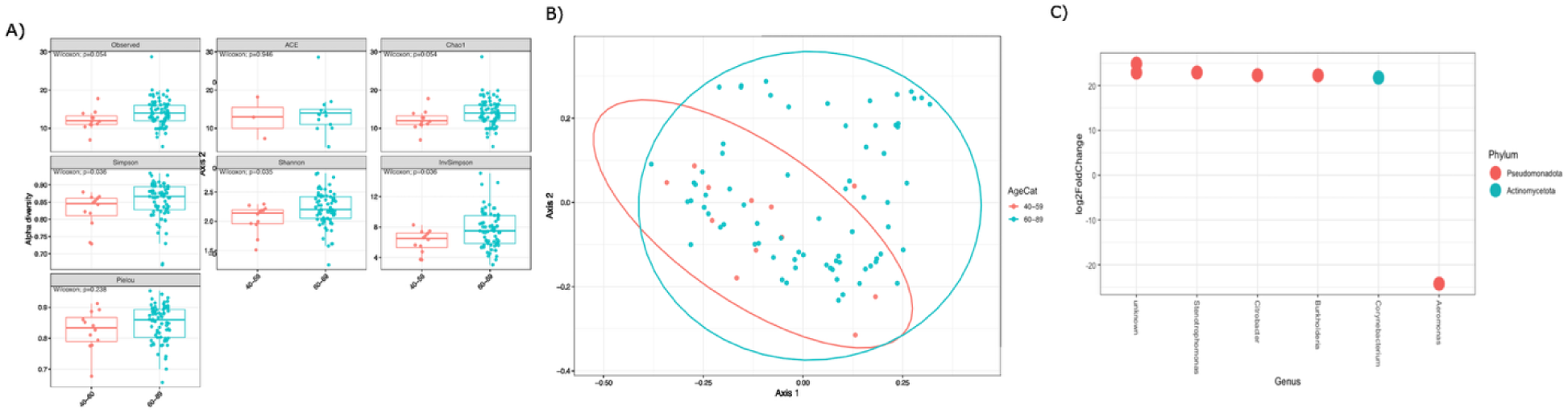
Comparison of microbial diversity in biopsy samples dichotomized by age (40-59 vs 60-89). (A) Comparisons of diversity by richness (Observed, Abundance-based coverage estimator [ACE] and Chao1), by evenness (Pielou) and by composites of richness, evenness, and abundance (Simpson, Shannon, Inverse Simpson), revealing significant differences in alpha diversity by Chao1, Simpson, Shannon, and Inverse Simpson. (B) Principal Coordinates Analysis (PCoA) based on the Bray-Curtis dissimilarity index, revealing significant differences in beta diversity (PERMANOVA p=0.053). (C) DESeq2 Analysis revealing enriched species in different age groups. Positive Log_2_FoldChange enriched in old (60-89); negative Log_2_FoldChange enriched in young (40-59).

When analyzing patient co-morbidities, we observed significant differences in the microbiome of patients with pre-operative atrial fibrillation (AFib), anti-coagulant use, and Chronic Obstructive Pulmonary Disease (COPD). For all three variables, significant differences were observed with the airway swabs. Participants diagnosed with AFib exhibited a significantly less diverse microbiome by overall alpha diversity indices measures (Simpson, Shannon, and Inverse Simpson; Kruskal-Wallis p=0.012, 0.018, and 0.012, respectively), primarily due to decreased richness (Observed and Chao1; Kruskal-Wallis p=0.030 for both; **Figure 3A**). Beta diversity analysis indicated a significant difference in composition (Bray-Curtis PERMANOVA p=0.01 and Jaccard PERMANOVA p=0.028; **Figure 3B**) with DESeq2 analysis revealing 6 ASVs significantly depleted in participants diagnosed with AFib: ASV11 (*Tropheryma*), ASV38 (*Comomonas*), ASV52 (*Pseudomonas*), ASV54 (Symbiopectobacterium) ASV65 (*Escherichia*), and ASV72 (*Ralstonia*; **Figure 3C**). Participants using anti-coagulants exhibited a significantly less diverse swab microbiome by one overall alpha diversity measure (Shannon; Kruskal-Wallis p=0.041) with two other measures trending towards the same result (Simpson and Inverse Simpson; Kruskal-Wallis p=0.072 for both), primarily due to decreased richness (Observed and Chao1; Kruskal-Wallis p=0.022 for both; **Figure 3D**). However, beta diversity revealed no significant difference in composition (PERMANOVA p>0.05). Finally, participants diagnosed with COPD also exhibited a significantly less diverse swab microbiome determined by alpha diversity indices of richness (Observed and Chao1; Kruskal-Wallis p=0.048 for both; **Figure 3E**). However, beta diversity revealed no significant difference in composition (PERMANOVA p>0.05).

**Figure 3.**
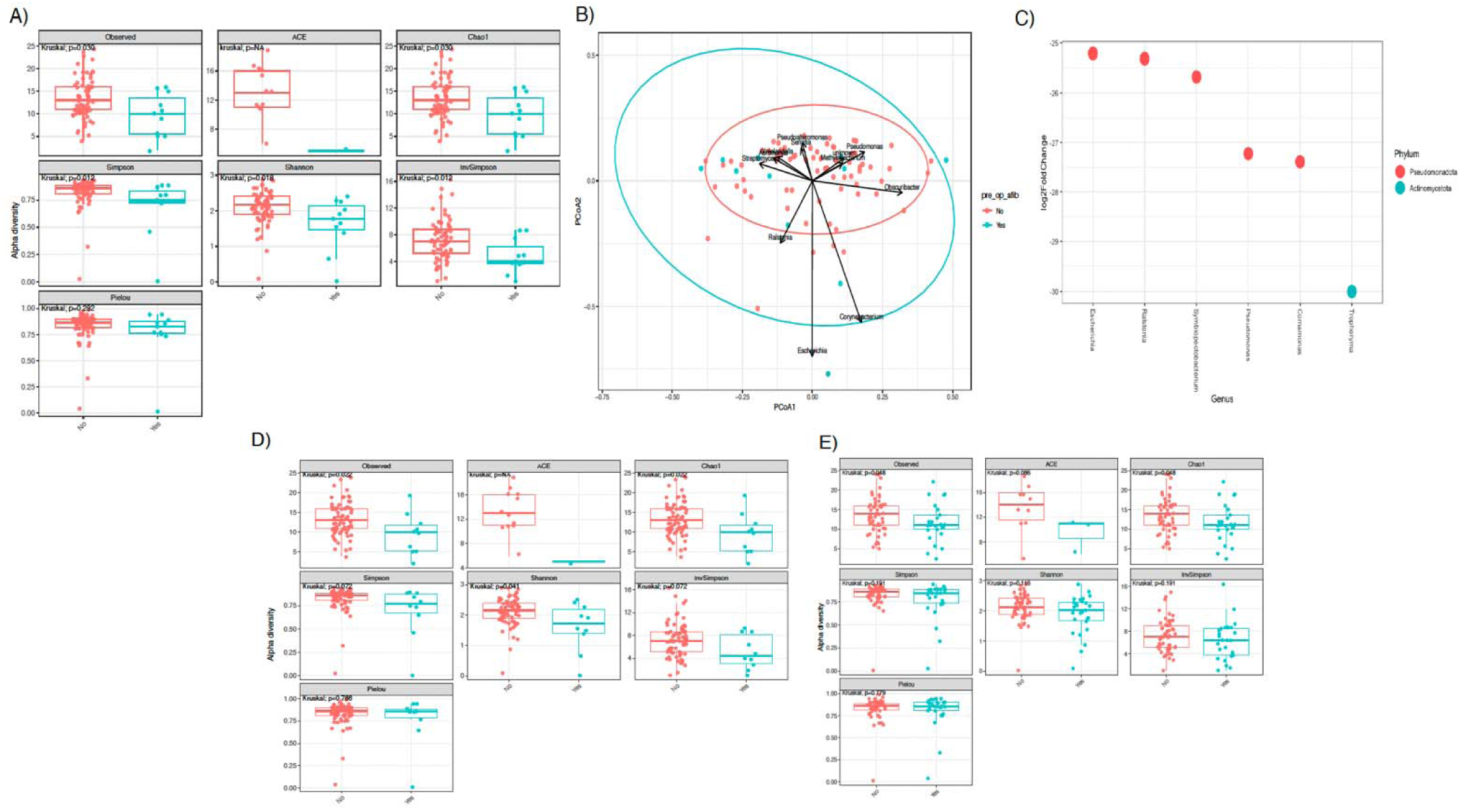
Comparison of microbial diversity across patient comorbidities. (A) Comparison of alpha diversity in patients with and without atrial fibrillation by richness (Observed, Abundance-based coverage estimator [ACE] and Chao1), by evenness (Pielou) and by composites of richness, evenness, and abundance (Simpson, Shannon, Inverse Simpson), revealing significant differences in alpha diversity by Simpson, Shannon, Inverse Simpson and richness (Observed and Chao 1). (B) Biplot of Principal Coordinates Analysis (PCoA) scores based on the Bray-Curtis dissimilarity index, revealing significant differences in beta diversity for patients with and without atrial fibrillation (Bray-Curtis PERMANOVA p=0.010; Jaccard PERMANOVA p=0.028). (C) DESeq2 Analysis revealing depleted species in patients with atrial fibrillation. Negative Log_2_FoldChange enriched in patients without atrial fibrillation compared to those with atrial fibrillation. (D) Comparison of alpha diversity in patients using and not using anticoagulants by richness, evenness, and composite measures, revealing significant differences in alpha diversity by Shannon (composite measure), Observed and Chao1 (richness). (E) Comparisons of diversity in patients with and without chronic obstructive pulmonary disease by richness, evenness, and composites measures, revealing significant differences in alpha diversity by richness (Observed and Chao1).

We then compared microbiome composition by lobe. For both sampling types, we constructed a histogram presenting the top 15 genera by relative abundance (**Figure 4A**). Whereas the parenchyma and airway microbiomes of the lobes did not differ by alpha diversity (**Supplemental Figures 4A-B**), the parenchymal microbiomes differed significantly by beta diversity (Bray PERMANOVA p=0.014; **Figure 4B**). In contrast, the airway microbiomes did not (**Supplemental Figure 4C**).

**Figure 4.**
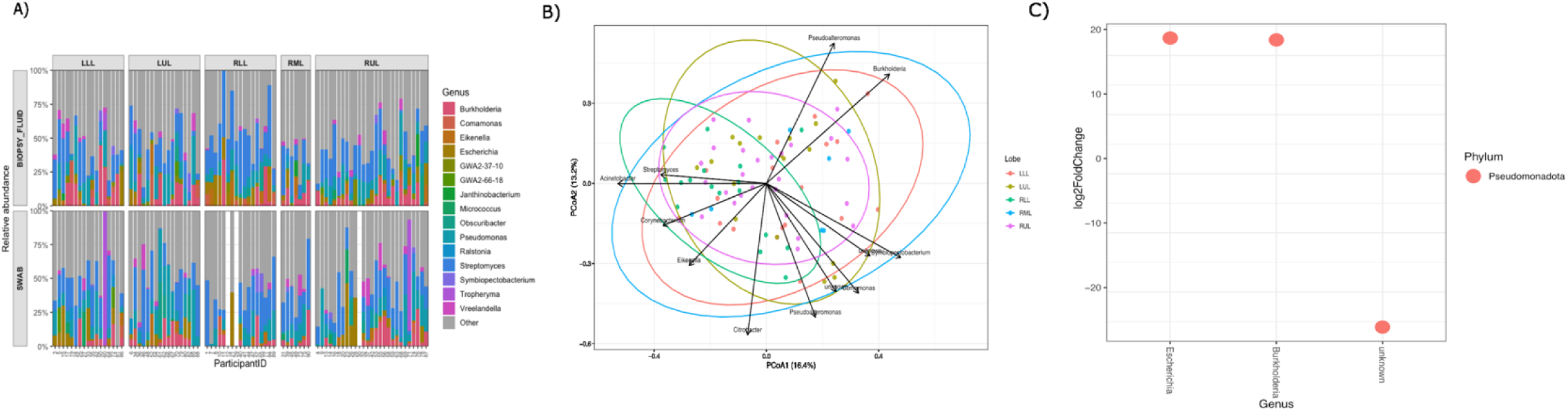
Comparison of microbial diversity by lobe. (A) Relative abundance of the top 15 species found in each of the five lobes; all other genera are included in “other.” (B) Biplot of Principal Coordinates Analysis (PCoA) scores based on the Bray-Curtis dissimilarity index, revealing significant differences in beta diversity (PERMANOVA p=0.014). (C) DESeq2 Analysis demonstrating enriched species in upper versus lower lobes. Positive Log_2_FoldChange enriched in upper lobes; negative Log_2_FoldChange enriched in lower lobes.

Finally, we compared left versus right lobes, and upper versus lower lobes. There were no significant differences in alpha diversity between left and right lobes (PERMANOVA p>0.05) or upper versus lower lobes. However, beta diversity analysis of parenchyma samples indicated a trend towards significance for upper versus lower lobes (PERMANOVA p=0.08). DESeq2 analysis indicated enrichment in the upper lobes of ASV40 (*Escherichia*) and ASV65 (*Burkholderia*) and enrichment in the lower lobes of ASV75 (unknown member of Rhizobiaceae family; p<0.001; **Figure 4C**).

## Discussion

In this prospective observational study of patients undergoing lung resection, we collected samples by two distinct methods (parenchymal biopsy and airway swabs). We found (1) the microbiome varies significantly between the parenchyma and the airway, (2) the microbiome composition varies across lobes of the lung, and (3) large variation exists between patients and their respective demographics and comorbidities. These results help explain previous diverse findings and may inform appropriate future sampling techniques, characterization, and analysis.

Current lung microbiome research is flawed by inconsistent sampling methods, including mixtures of BAL, sputum samples, bronchial brushing, and direct lung tissue sampling, making comparison and interpretation difficult between studies.^19^ Results have varied with diverse microbes reported in different studies and patient populations, highlighting the need for optimized sampling methods. Our findings concerning two different sampling methods provide some clarity to previously mixed results.^19^ Our beta diversity analysis revealed a significant difference in lung microbiome composition based on sampling method, with ASVs from three different genera differing between parenchyma and airway. Prior study results revealed higher bacterial densities in upper versus lower airways,^11,20^ but our results now also show differences in composition of the microbiome within these airways. Since ASV numbers are ranked increasingly by relative abundance, it’s likely that enriched ASVs are not rare members of these communities. Ultimately, our results indicate that upper and lower airways require separate sampling and cannot simply be lumped into one group.

There are several possible reasons why microbial composition may vary in different areas of the lung. For one, different bacteria thrive on different nutrients and environmental factors secondary to differences in metabolic pathways. In the lung, there may be differences in microenvironmental factors between parenchyma compared to large airways, such as pH, ventilation, and humidity.^21–23^ These differences may also play a role in development of different respiratory diseases and tumor progression.^23,24^ However, it is unclear if some of these diseases, and particularly cancer, lead to changes in the microbiome or vice versa. Regardless, study of these interactions could lead to improved prognostication, prediction, and ultimately, treatment of these conditions.

In comparisons between parenchymal and airway samples, ASVs from the genera *Pseudomonas* and *Corynebacterium* were enriched in biopsy tissue, while an ASV associated with the genus GWA2-37-10 was enriched in the swab samples. Both *Pseudomonas* and *Corynebacterium* have been noted to be involved in human cancer development and harbor potential immunomodulatory mechanisms.^25–27^ On the contrary, there is a scarcity of literature regarding the “ultrasmall” bacterial genus GWA2-37-10.^28^ Some studies suggest associations with dysbiosis, cancer, and inflammation, but the causality behind these associations remains unclear.^28,29^ Thus, further work is needed to determine how these genera impact cancer progression and what influences their enrichment. However, our results indicate that these microbes may only be enriched in the most distal airways/parenchyma, so proximal sampling strategies (*i.e.*, BAL) may not be sufficient.

Similarly, there were notable differences in beta diversity among lobes of the lung when comparing parenchymal samples between all five lobes. Of note, while we measured no difference in alpha or beta diversity based on laterality, we did note a near-significant trend in beta diversity between upper and lower lobes. This finding may be more intuitive as we know aeration varies throughout the lung and these differences can lead to certain disease patterns. For example, *Mycobacterium tuberculosis* is largely found in the apex, theoretically secondary to decreased ventilation and arterial flow in this area.^30^ Likewise, lobe and tumor location may differentially alter aeration and blood flow, leading to changes in the microbiome of these regions. Lung cancers also tend to favor the upper lobes, likely secondary to increased exposure to inhaled toxins.^31^ Prior studies have indicated similar trends,^32^ so our results confirm and expand on these findings by identifying species that are enriched in certain areas of the lung. In our study, the upper lobes were enriched with ASVs associated with the genera *Escherichia* and *Burkholderia,* while the lower lobes were enriched with a member of the family Rhizobiaceae. In prior studies, *Escherichia* has been linked to the promotion of lung cancer,^33^ and Rhizobiaceae has been linked to tumor progression in other disease sites (i.e., liver cancer).^34^ Therefore, it’s possible that these microbes could represent a clinically meaningful impact on cancer prediction and progression.

Further sub-analyses of patient demographics uncovered significant differences in parenchyma with respect to age, with older participants exhibiting a more diverse microbiome enriched with ASVs from the phylum Pseudomonadota and the genera *Corynebacterium, Citrobacter*, *Burkholderia*, and *Stenotrophomonas*. Prior studies have reported differences in lung microbiome across different age groups, but many of these samples were taken in the context of advanced pulmonary disease, such as cystic fibrosis, which presents a significant confounding factor.^35^ Thus, in these studies, it is unclear whether a true association with advanced age exists or if underlying diseases were confounding. In the present investigation, we studied a more homogenous group of patients, all obtaining care at the same hospital for the same indication, revealing that the lung microbiome may be differentially shaped by environmental exposures over a patient’s lifetime.

However, one important negative finding in our results was the lack of differences seen in the microbiome of smokers and nonsmokers, which was the opposite of what we had anticipated. It is widely suspected that smoking influences the lung microbiome, but existing literature on this topic is mixed. Several prior studies have shown no difference in the lung microbiome by smoking status, but have shown changes to oropharyngeal flora.^36,37^ Meanwhile, others have identified specific taxa (*e.g., Acidovorax*) that are enriched in smokers.^24^ One recent study showed that the lung microbiome can shift for weeks to years after smoking cessation.^38^ Our cohort consisted of 75.6% smokers, with a mean of 36.2 pack-years (range 1-102). However, date of smoking cessation was unknown, so it is possible that the heterogenous nature of smoking behavior in our cohort resulted in the null finding.

On the contrary, the lung microbiome did vary with underlying diseases and medical conditions, such as COPD and atrial fibrillation. It may not be surprising that intrinsic lung conditions, such as chronic obstructive pulmonary disease are associated with a unique microbiome, given chronic changes in expiratory forces, ventilation, and prior exposures that may have contributed to the development of COPD.^39^ However, this is an important association to note in this study as it demonstrates one of several reasons why the lung microbiome may differ between patients. Prior studies have also shown differences in patients with severe COPD compared to those without COPD,^40^ and our study confirms this finding. In addition, it highlights the difference in impact between temporary exposures (i.e., smoking) and chronic diseases (i.e., COPD) even if the two may be related.

Furthermore, our findings regarding differences in patients with atrial fibrillation and those treated with anticoagulants may be less intuitive, but similar results have been shown in the gut microbiome.^41–43^ To our knowledge, this is the first time it has been demonstrated in the lung, showing that these diseases and medications may have systemic interactions with the body’s microbiome. It’s postulated that there may be an underlying gut-immune-heart axis whereby gut dysbiosis contributes to chronic inflammation, oxidative stress, fibrosis, and remodeling of atrial tissue.^41^ Of note, difference in the microbiome of patients with atrial fibrillation was only seen in airway samples and not in the parenchyma, so there may be differential impacts of these underlying diseases based on location within the lung and associated blood supply. Regardless, our study now provides further evidence towards the theorized bidirectional influence between microbiota, lung and heart health.^44,45^

Other demographic and clinical factors, including sex, race, ethnicity, body mass index, and steroid use showed no differences in the lung microbiome. Existing literature reports mixed results on these factors, likely owing to small sample sizes and significant heterogeneity between patients.^11,36,46^ For example, a systematic review on the effects of corticosteroids on the respiratory microbiome demonstrated significant heterogeneity in findings, but also highlighted the heterogeneity in sampling methods across studies, which our study aims to improve.^47^ Thus, although we saw no differences, large-scale studies using consistent sampling methods should be used to further elucidate these findings.

This study has several strengths. First, all samples were obtained in a sterile setting within a single institution and kept cold until addition of a nucleic acid preservative to minimize potential for contamination. We included negative controls and subjected sequence data to a robust bioinformatic decontamination procedure. A quality control analysis revealed no differences associated with the surgeon performing the operation, which could suggest contamination, but fortunately this was not the case. Furthermore, all samples were sequenced simultaneously to reduce batch effect.

Our study has a few limitations. First, we could not account for precise distance between tumor tissue and biopsy samples. Prior studies have indicated that tumor tissue itself may harbor its own microbiome,^20^ so this could alter results. However, all samples were taken from the same lobe as the tumor, and consistent sampling methods were employed. Second, our patient sample was limited to a single institution for a single indication that may be skewed toward certain demographics (*i.e.*, white, elderly, smokers), which limits generalizability.

However, this also made for a more homogenous patient population, which improved our ability to compare groups. Finally, all patients received standard surgical antimicrobial prophylaxis at initiation of their operations. Theoretically, this could alter the lung microbiome, but all samples were taken within hours, so sequencing would still identify bacterial remnants.

In conclusion, appropriate sampling methods for the lung microbiome are of utmost importance. Apparent differences exist between the microbiome of airways and parenchyma, which may differentially impact study results and patient outcomes. The present study adds substantial data to the literature and may serve as a guide for future standardized lung microbiome analysis.

## Data Availability

Data is accessible at NCBI BioProject: ID#PRJNA1428797, entitled "Pulmonary Oncogenesis Microbiome Following Surgery."

## Acknowledgements

We would like to thank the surgeons who collected specimens, including Drs. Julia Coughlin, James Lubawksi, Wissam Raad, and Wickii Vigneswaran. We would also like to thank clinical/research personnel who assisted with sample collection and processing, including Raymond Verm, Madison Lozanoski, Mohamed Gadelkarim, Ayham Odeh, Bilal Odeh, Joseph Zywiciel Jr, Omar Zahra, and Germaine Harvey. Finally, we are deeply grateful to all the patients whose participation made this study possible.

## Author Contributions (CRediT)

**Alexander Pohlman**: methodology, validation, investigation, data curation, visualization, writing – original draft, writing – review and editing. **Andrew Marten**: methodology, validation, data curation, writing – original draft, writing – review and editing. **Melline Fontes Noronha**: Methodology, software, validation, formal analysis, data curation, writing – original draft, writing – review and editing, visualization. **Mark Khemmani**: Methodology, validation, investigation, writing – original draft, writing – review and editing. **Alan Wolfe**: Conceptualization, methodology, validation, resources, project administration, supervision, writing – original draft, writing – reviews and editing, funding acquisition. **Zaid Abdelsattar**: Conceptualization, methodology, validation, resources, project administration, supervision, writing – original draft, writing – reviews and editing, funding acquisition.

