## Supplemental Material for "Sampling of the Lung Microbiome in Patients Undergoing Lung Resection"

**Supplemental Table 1**. Definitions for measures of microbial diversity.

| **Term** | **Definition** |
| --- | --- |
| Alpha Diversity | Measures taxonomic composition within a given sample |
| Observed, Chao1, and ACE | Measures richness (the number of unique taxa per sample). More diverse samples have larger values |
| Pielou | Measures evenness and refers to the distribution of taxa. More evenly distributed samples have larger values |
| Shannon, Simpson, and Inverse Simpson | Composite indices that include richness, evenness, and abundance in their calculations |
| Beta Diversity | Between sample diversity |

*ACE – abundance-based coverage estimator

**Supplemental Figure 1**. Overview of DNA extraction and sequencing process. **Created in BioRender. Pohlman, A. (2026) https://BioRender.com/pbi37mm*

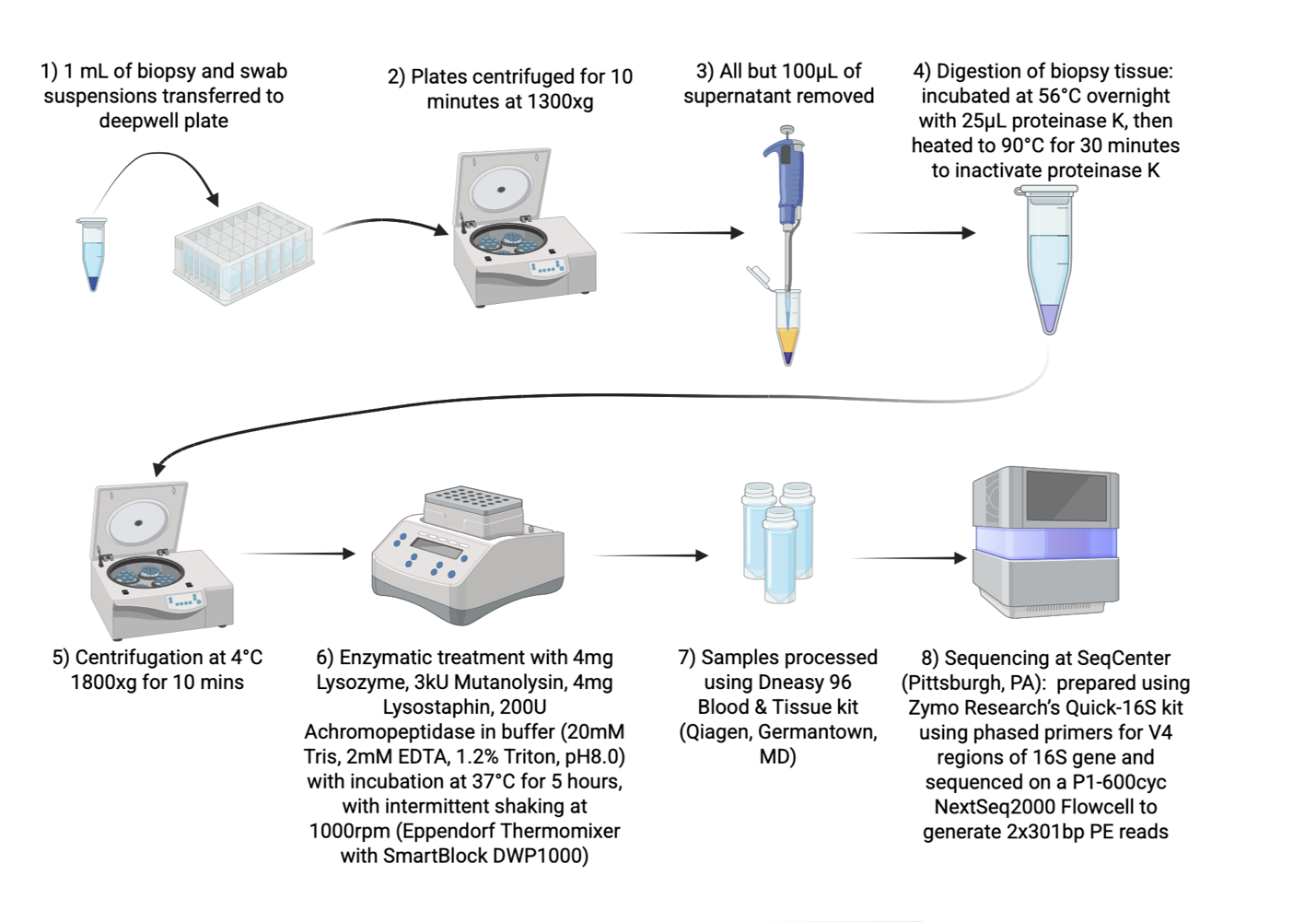
**Supplemental Figure 2.** Comparison of microbial diversity of participants by sampling method: biopsy and swab. (A) Principal Coordinates Analysis (PCoA) based on the Bray-Curtis dissimilarity index, revealing significant differences by participant biopsy sample (PERMANOVA p<0.001). (B) Principal Coordinates Analysis (PCoA) based on the Bray-Curtis dissimilarity index, revealing significant differences by participant swab sample (PERMANOVA p<0.001).

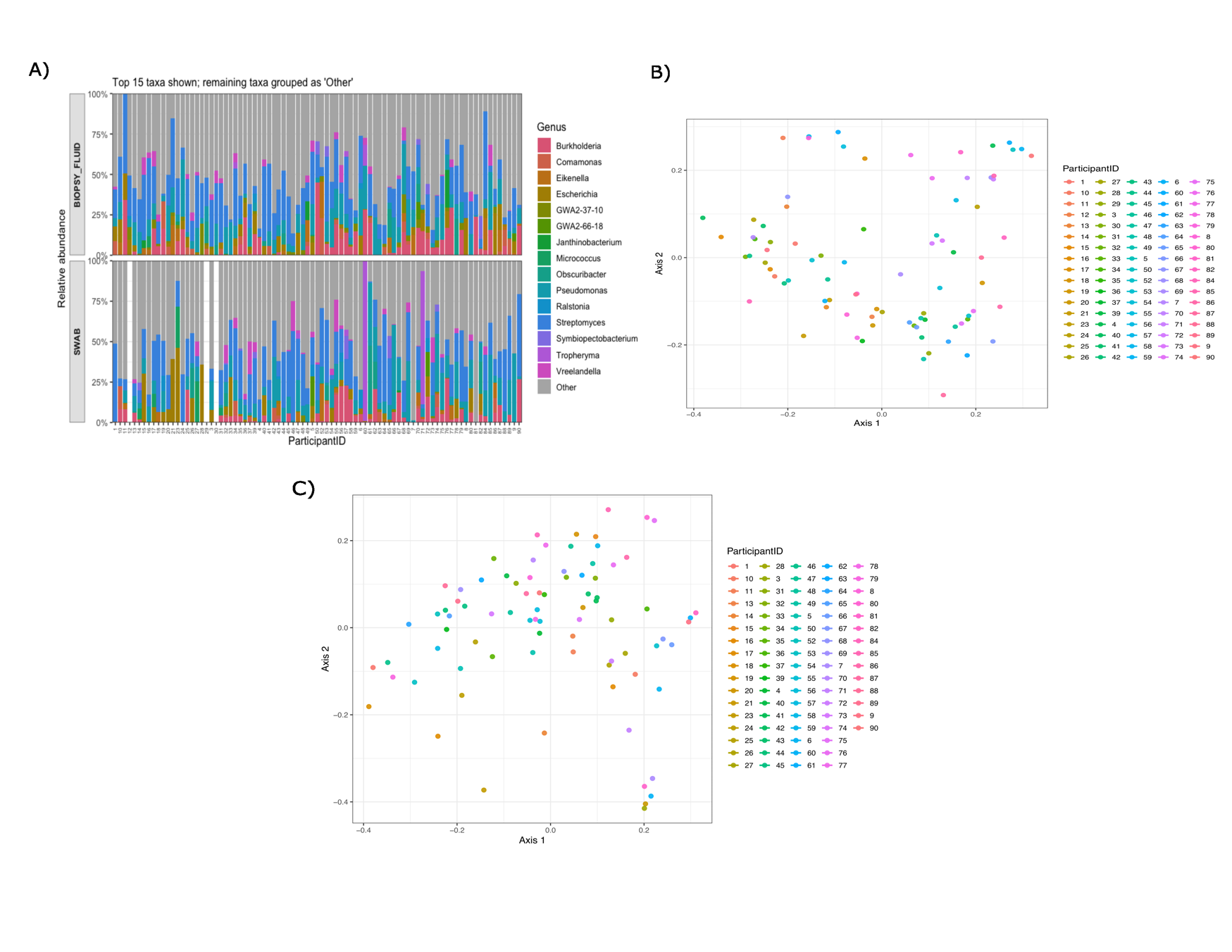

A)

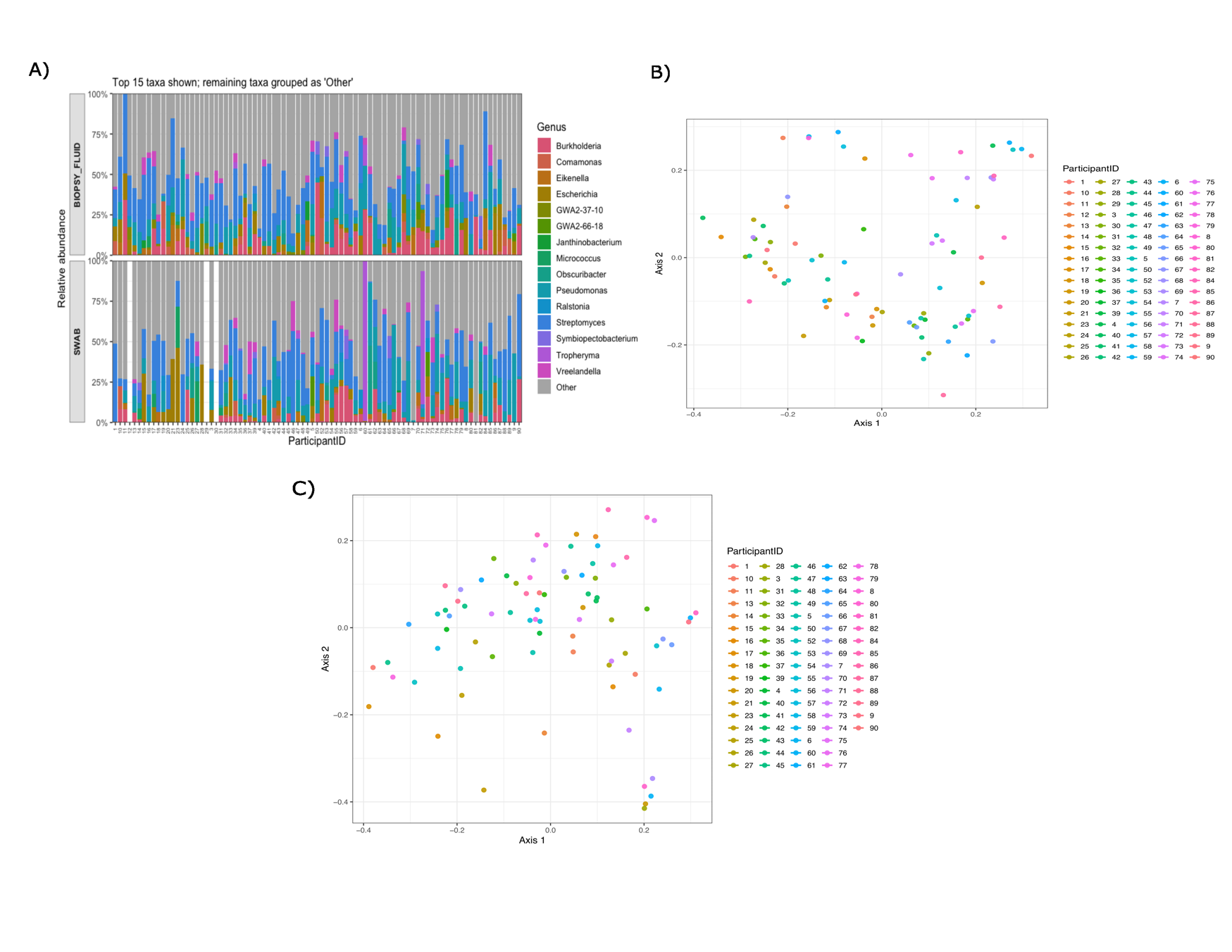

B)

**Supplemental Figure 3.** Comparison of microbial diversity in swabs dichotomized by age range (40-59 vs 60-89). (A) Comparisons of alpha diversity by richness (Observed, Abundance-based conversion estimator [ACE] and Chao1), by evenness (Pielou) and by a composite of richness, evenness, and relative abundance (Simpson, Shannon, and Inverse Simpson), all showing no significant differences. (B) Principal Coordinates Analysis (PCoA) based on the Bray-Curtis dissimilarity index, revealing no significant differences in beta diversity (PERMANOVA p>0.05).

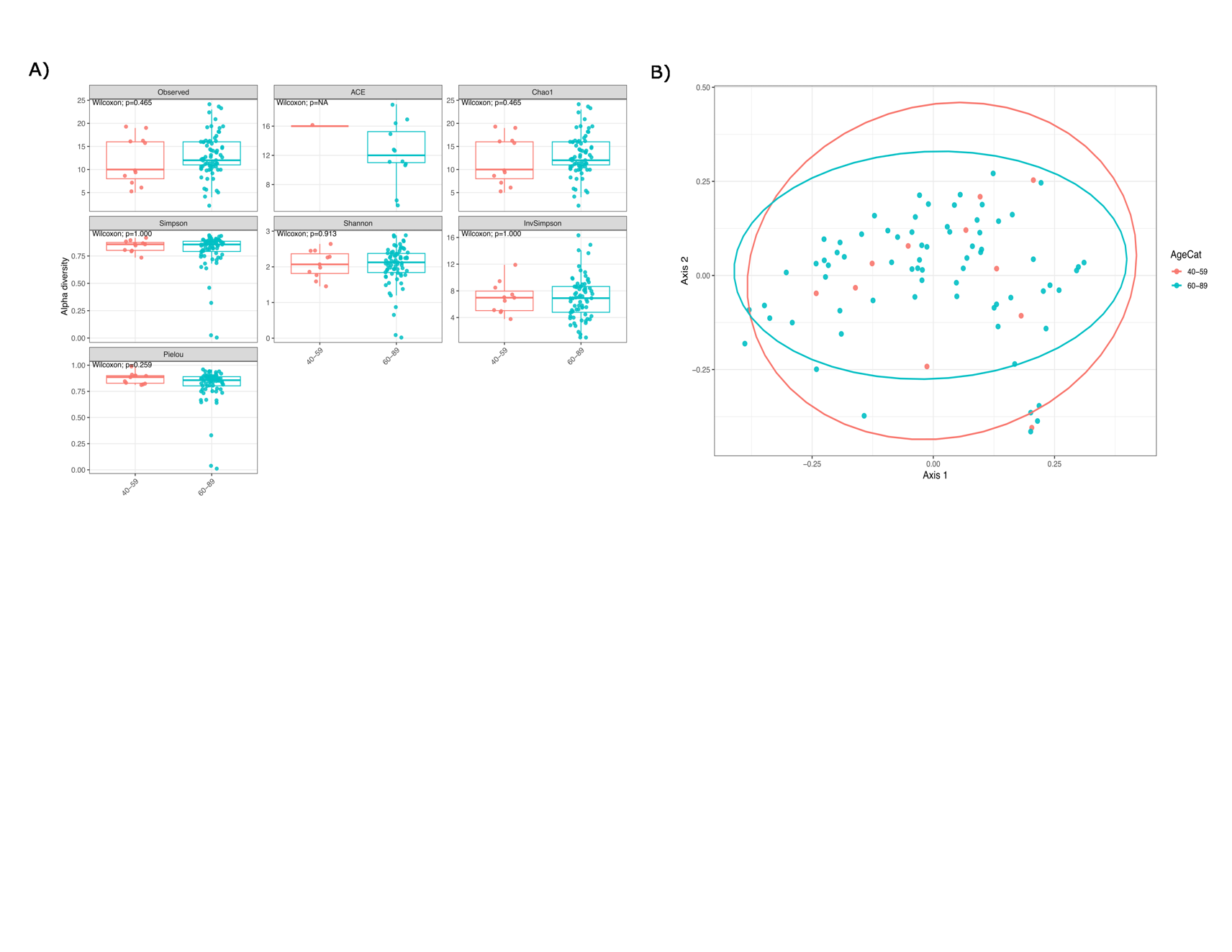

**Supplemental Figure 4.** Comparison of microbial diversity by anatomic location (lobe of the lung). (A and B) Comparisons of diversity by richness (Observed, Abundance-based conversion estimator [ACE] and Chao1), by evenness (Pielou) and by a composite of richness, evenness, and abundance (Simpson, Shannon, and Inverse Simpson), revealing no significant differences. A, biopsy samples; B, swabs. (C) A biplot of Principal Coordinates Analysis (PCoA) scores based on the Bray-Curtis dissimilarity index, revealing no significant differences in beta diversity of swab samples (PERMANOVA p>0.05).

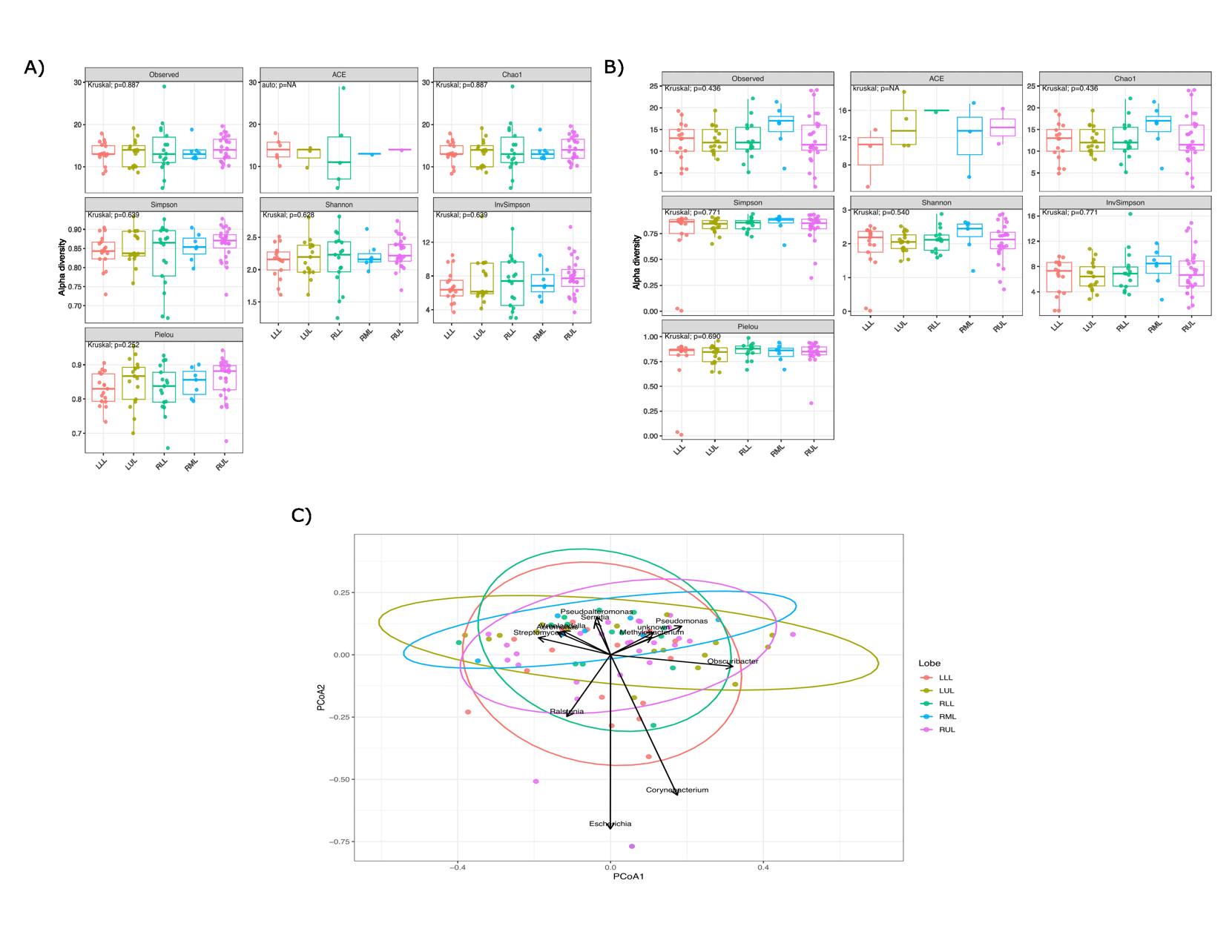
